# Daily Experiences of Urinary and Fecal Incontinence in Young Adults with Spina Bifida: Preliminary Results from an Ecological Momentary Assessment Study

**DOI:** 10.1101/2025.01.10.24313751

**Authors:** Devon J. Hensel, Audrey I. Young, Konrad M. Szymanski

**Author notes:** **Corresponding author:** Devon J. Hensel, 410 W. 10th Street, Room 1001, Indianapolis, IN 46202. **Funding**: This project was funded by a National Institute of Digestive and Kidney Disorders (NIDDK) (R21DK121355) to Drs. Hensel and Szymanski. The work presented here reflects the viewpoints of the authors and not necessarily those of NIDDK. **Data Availability:** Raw data files and codebooks are stored with the Open Science Framework (https://osf.io/gefqx/).

## Abstract

**Background:** Urinary (UI) and fecal (FI) incontinence are prevalent secondary chronic conditions among young adults with spina bifida (YASB). UI and FI decrease daily functioning for YASB, but no research has prospectively examined characteristics of UI and FI among YASB. We used ecological momentary assessment (EMA) over 30 days to describe the prevalence, episode-specific characteristics and negativity associated with UI and FI among a cohort of YASB.

**Method:** Data were collected as part of a larger 30-day EMA study prospectively examining the daily prevalence and context of UI and FI in adults with SB. We drew an analytic sample of young adults (YASB) participants aged 18-27 years (N=23 [26.1% of all study-participants [N=88];). Participants completed an end-of-day EMA tracking the frequency, dry intervals, volume, activity avoidance, management, positive and negative mood, current UI (UIA) or FI (FIA) anxiety, past UIA and FIA and past number of UI or FI events.

**Results:** YASB contributed a 643 daily EMAs. Nearly 60% (370/643) of all daily entries was associated with a general report of incontinence (UI: 54.1% [348/643]; FI: 20.8% [134/643]). Prevalence, characteristics and negativity associated with UI and FI varied significantly from day-to-day. Higher UI frequency, shorter dry intervals, greater UI volume, needing management help, avoiding activities because of UI, higher past median UIA, higher negative mood and fewer past UI events were associated with higher daily UIA. Shorter FI dry intervals, higher past median FIA, higher negative mood and fewer past FI events were associated with higher daily FIA.

**Discussion:** Day-to-day experiences of UI and FI vary among adults with SB across multiple dimensions. Negativity about incontinence when it occurs varies not only based on individual- and episode-specific characteristics, but also on incontinence in the preceding days. Operationalizing these insights into potential clinical interventions warrants further investigation.

**Discussion and Implications:** Young people with SB (YASB) experience day-to-day differences in the characteristics of urinary (UI) and fecal (FI) incontinence (e.g. frequency, self-management) events and the way they felt (e.g. affect) about UI and FI. The unique relationship of these factors to UI and FI anxiety suggest novel potential points of intervention.

## Background

Urinary (UI) and fecal (FI) incontinence are common secondary health conditions among young adults with spina bifida (YASB). Large scale studies report past month UI prevalence between 50% and 75% and past month FI prevalence between 41% and 55%.^1–4^ Many YASB report both UI and FI in the past month.^5^ UI and FI can interfere with YASB’s ability to participate in daily tasks (e.g. managing peer/romantic relationships, finishing education, finding employment) that are necessary for longer-term independent living.^6–8^

Clinical or research evaluation of UI or FI “impact” typically relies on questionnaires that ask YASB to retrospectively recall a handful of primarily objective (e.g., mild or severe leaking, number of dry intervals) incontinence symptoms across what is often a long time block (e.g., “in the past month” or “since the last clinic visit”).^9^ Although these approaches are common because it is difficulty for a clinician or researcher to directly observe incontinence in a person’s daily life, they can create challenges with the information provided. Longer recall periods can decrease YASB ability to call accuracy past incontinence details,^10^ or they may underreport UI or FI out of concern privacy or embarrassment.^11^ Moreover, summarizing multiple data points a single data point (e.g., aggregating 30 daily UI or FI experiences into a single “month” focused question) reduces the ability to understand how different aspects of UI and FI – like frequency, amount or activity interference – may look differently from one day to the next.^12–14^ Both clinical experience and recent research^5,15–17^ suggest that daily life with incontinence varies not only from person to person, but also within the same person over time. If a goal is to capture “clinically significant” UI and FI for the purpose of clinical decision making, incontinence instruments must have the accuracy and the sensitivity to fully screen any daily UI or FI episode for different potential problems.^10,18^ A variety of factors – some perhaps even different across several incontinence events – can uniquely impact how YASB experience UI and FI. Capture of these variations is therefore an essential part of personalized continence care, as therapies can be directly designed and adapted from YASB’ own “real life” experiences.^19^

In the current paper, we used ecological momentary assessment (EMA) to collect these needed data. EMA is a broad class of prospective – or forward looking – data collection approaches intended to capture snapshots of people’s daily lives through brief-but-repeated assessments in their “natural” or “natural” environments.^20^ Data are typically collected using the functional capabilities of web-enabled electronic devices, and participants provide data on a study-specific time schedules (for example, once/day or following a specific health event; for an excellent review, see Hufford et. al, 2007 ^21^) usually over weeks or months.^22,23^ In the context of incontinence among YASB, a given EMA provides an “in the moment” view of a UI or FI event by asking YASB to provide an account of both the symptoms of that day’s UI and FI (e.g., frequency, amount, dry intervals) and the context in which the UI or FI events occurred (e.g., feeling negatively about incontinence, any activity avoidance, self-management).^24^ Over time, the collective of these data points constructs a portrait of “typical” or “everyday” incontinence as it is “lived by” YASB themselves.^25^ With enough data, it is possible to evaluate what life looks on days when UI and FI do not occur, as well as how the relationship between UI or FI characteristics and context may change over time.^26^ Such data are important for optimizing the chance that interventions based on this knowledge will be effective when employed in people’s daily lives.^27^ A more complete review of the advantages of EMA use in the SB population is available in Hensel et. al, 2023.^9^

We use these data to address important unknowns about incontinence for YASB. One question is – *how often* do either UI and FI occur, and when they do, what does the *characteristics* of a “typical” event look like? Do either the prevalence or the characteristics vary from day-to-day? A second question is – does the way in which a YASB experience UI or FI impact how they feel about the incontinence event? In other words, do some characteristics, but not others, increase the anxiety YASB feel about UI or FI? These very basic questions remain largely unaddressed in the extant SB literature. As a result, several community calls have been made for research than can more clearly document what it is like for YASB to “live with” incontinence.^28^

Accordingly, the primary objectives of our paper were to:

1. Describe the daily *prevalence* (e.g. how often) and *characteristics* (e.g., amount, dry intervals, affect or activity avoidance) of UI and FI among YASB (Objective 1a), and examine the extent to which either of them vary from day-to-day (Objective 1b).
2. Evaluate the preliminary correlation of prevalence and characteristics with any *anxiety* YASB feel about UI or FI.

## Methods

### Data and Participants

Data were drawn from a larger 30-day study (R21DK121355) that examined the feasibility of using ecological momentary assessment (EMA) to describe daily incontinence experiences among adults with SB. Participants were recruited through social media (e.g. researcher or SB association posts), clinical referral or through completed participant referral. Study eligibility requirements included being 18 years or older, having SB, having any UI and/or FI in the past four weeks, having English language literacy and having normal to mildly impaired cognitive development. As part of this larger study, participants completed 30 end-of-day EMAs that assessed UI and/or FI prevalence and episode-specific characteristics, any UI or FI negativity and daily affect. The larger project was approved by the Institutional Review Board of Indiana University (#1907916729), and achieved excellent retention (97.5%), EMA completion (95.8%), and data accuracy (98.6%) and acceptability (>90% enjoyed the study, found protocol easy, felt supported, would participate again).^9,18^

For the current paper, we drew an analytic sample of AYSB participants aged 18-27 years (N=23 [26.1% of all study-participants [N=88];). These AYSB were mostly female (78.3%), White (91.3%) and heterosexual (82.6%) and had a median educational completion of some college. Half reported their relationship status to be single and about a third were working part- or full-time. More than half of AYASB reported having a ventriculoperitoneal shunt (VPS), and about 40% were community ambulators. A third (39.1%) lived independently while most (60.1%) lived with one or more parents. All reported some kind of incontinence in the four weeks prior to the study (17.3%: UI only, 13.0% reported FI only; 70% both UI and FI. In addition, all reported some kind of incontinence during the 30-day diary period: 21.7% (5/23) of YASB reported UI only in the diaries, 4.3% (1/23) reported FI only, and 73.9% (17/23) reported experiencing both UI and FI during the diary period (Supplemental Table). Additional person-level analyses are described in the Supplement.

### Measures

#### Daily UI and FI Prevalence

Incontinence frequency (original item: once, twice, three to five times, six or more times; item dichotomized for this work based on low frequencies at the higher end: once vs. two or more times).

#### Daily UI and FI Symptoms

Variables were: volume (small [a little bit], medium, large [a lot]) and length of dry intervals between UI or FI (never dry, less than four hours, more than four hours).

#### Daily UI and FI Context

Variables were: avoided any activities due to incontinence (no/yes) and incontinence management (fully on my own, needed some help, needed a lot of help).

#### Daily Affect

We included positive mood (“happy” and “friendly”) and negative mood (“angry” and “irritable”) (PANAS; both additive indices of the two, 5-point Likert items [not at all to extremely]).^29^

#### Daily Incontinence Anxiety

We used a single Likert- type items that asked: “To what extent did you feel anxious about [incontinence type] today?” (not at all or very slightly, a little, moderately, extremely).

#### Past incontinence experiences

We included two time lagged variables: past incontinence frequency (summed number of UI or FI events through the prior day) and past UI anxiety or FI anxiety (running median; scale described above).

### Statistical Analysis

All analyses described below were performed in Stata (v. 18.0).^30^

#### Objective 1a – Description of UI or FI Prevalence and Characteristics

We used descriptive statistics to characterize the daily distribution of UI and FI variables.

#### Objective 1b – Day-to-day Variation in UI or FI Prevalence and Characteristics

We used intraclass correlation coefficients (ICC) to descriptively quantify the amount of relative between- or within-AYASB variability in outcomes. Larger ICCs indicate more differences between-persons, smaller ICCs suggest more within-person differences.^31^ Because this was an exploratory study, we did not have any *apriori* hypotheses regarding the magnitude of any ICC and describe them for context only.

#### Objective 2 – Preliminary Daily Association Between UI and FI Prevalence and Characteristics with UI (UIA) or FI (FIA) Anxiety

We used Spearman rank correlation coefficient to examine the preliminary associations between prevalence and characteristics with UIA and FIA.^32^

## Results

### UI and FI Daily Summary

YASB contributed a total of 643 daily entries during their time in the study, representing 93.1% completion of the total entries expected (N=690). Nearly 60% (370/643) of all daily entries was associated with a general report of incontinence, meaning that YASB reported some type of incontinence on about *four out of every seven days in a “typical” week*. On these days, 63.7% (236/370) were only UI, 9.3% were FI only (22/370) and 30.2% (112/370) were both UI and FI. Neither the general occurrence of incontinence (*p*=0.826) nor the type of incontinence was more likely to occur on any given day of the week as compared to others (*p*=0.505).

### Objective 1a – Describe day-Level Variation of Prevalence and Characteristics of UI and FI

#### Urinary incontinence

As shown in Table 2, YASB reported any UI on slightly more than half of all days (54.12%: 348/643). In other words, YASB generally experienced any UI about *three out of every seven days in a “typical” week*. Sensitivity analyses suggest that reports of UI did not differ significantly by day of the week (*p*=0.272). Reporting UI did not change as a function of study participation progression (e.g., Day 1 vs. Day 13 vs. Day 29).^9^

**Table 1.** Characteristics of Young Adults with Spina Bifida (YASB).

| Characteristic | Percent (N) or Mean (SD); Median |
| --- | --- |
| Age | 22.9 (2.31); 23.0 |
| Birth-assigned sex (female) | 78.3 |
| Gender identity (female) | 78.3 |
| Sexual identity |  |
| Heterosexual | 82.6 |
| Gay or lesbian | 0 |
| Bisexual | 13 |
| Asexual | 4.32 |
| Something else | 0 |
| Race/ethnicity |  |
| White | 91.3 |
| Black | 8.7 |
| American Indian or Alaska Native | 0.0 |
| Asian | 0.0 |
| Other | 0.0 |
| Multiple races | 0.0 |
| Hispanic or Latino (yes) | 4.4 |
| Annual household income |  |
| <\$20,000 | 20.0 |
| \$20,000 to \$39,000 | 15.0 |
| \$40,000 to \$59,000 | 10.0 |
| \$60,000 to \$79,000 | 5.0 |
| \$80,000 to \$100,000 | 5.0 |
| \$100,000 or more | 15.0 |
| Unsure | 30.0 |
| Highest education level completed |  |
| Less than high school | 0.0 |
| High school degree or equivalent | 39.3 |
| Some college but no degree | 17.3 |
| Bachelor's degree | 34.7 |
| Graduate degree | 8.0 |
| Current work status |  |
| Working full time | 17.3 |
| Working part time | 26.1 |
| Unemployed and looking for work | 17.3 |
| Unemployed and not looking for work | 13.0 |
| On disability | 26.1 |
| Currently attending school (full or part times: yes) | 17.3 |
| Current relationship status |  |
| Single and not dating | 56.5 |
| Single and dating but not in a relationship | 8.7 |
| In a relationship, but not living together | 13.0 |
| In a relationship, living together (not married) | 8.7 |
| Married | 13.0 |
| Divorced | 0.0 |
| Widowed | 0.0 |
| Currently living independently (yes) | 39.3 |
| Ambulation |  |
| I move only in my wheelchair | 26.1 |
| I walk only during therapy. I use a wheelchair the rest of the time | 4.3 |
| I walk inside and outside (with or without a walker, braces or | 17.4 |
| crutches). I only use a wheelchair for long trips.<br>I walk inside and outside without any help | 26.1 |
| Have a ventriculoperitoneal shunt (VPS) (yes) | 60.8 |
| Any incontinence at enrollment (past four weeks: yes) | 100.0 |
| UI only | 17.4 |
| FI only | 13.0 |
| Both UI and FI | 69.6 |
| Catheterize bladder (yes) | 87.1 |
| Malone antegrade colonic enema (MACE) to empty bowels (yes) | 18.8 |

**Table 2.** Day-Level Information of Urinary and Fecal Incontinence among a Sample of Young Adults with Spina Bifida.

| Incontinence Daily Measure | Urinary Incontinence |  |  | Fecal Incontinence |  |  |
| --- | --- | --- | --- | --- | --- | --- |
|  | Percent or Mean (SD) and Median | ICC (95% CI) | Spearman Correlation with UI Anxiety | Percent or Mean (SD) and Median | ICC (95% CI) | Spearman Correlation with FI Anxiety |
| Frequency<br>Once (ref)<br>Two or more times | 44.40<br>55.60 | 0.86 (0.69 – 0.94) | 0.120*** | 82.09<br>11.19<br>6.72 | 0.50 (0.32 – 0.71) | 0.157*** |
| Dry Intervals<br>More than 4 hours (ref)<br>Less than 4 hours<br>Never dry | 31.32<br>54.60<br>14.08 | 0.75 (0.58 – 0.86) | -0.148*** | 54.89<br>35.34<br>9.77 | 0.43 (0.17 – 0.73) | 0.031 |
| Amount<br>Small (ref)<br>Medium<br>Large | 25.00<br>55.75<br>19.25 | 0.78 (0.62 – 0.89) | 0.216*** | 52.99<br>33.58<br>13.43 | 0.44 (0.21 – 0.71) | 0.269*** |
| Independent management (yes) | 89.30 | 0.96 (0.85 – 0.99) | 0.034 | 83.30 | 0.85 (0.64 – 0.95) | -0.131** |
| Avoided any activities (yes) | 7.76 | 0.89 (0.56 – 0.98) | 0.180*** | 20.10 | 0.43 (0.16 – 0.76) | 0.416*** |
| Number of past incontinence days (range: 0 – 29) | 8.15 (8.40); 5.00 | 0.32 (0.16 – 0.54) | 0.113 | 3.23 (5.16); 1.00 | 0.87 (0.74 – 0.94) | -0.114** |
| Median past incontinence anxiety (range: 1 – 5) | 3.95 (5.16); 4.00 | 0.92 (0.673 – 0.98) | 0.471*** | 3.21 (1.70); 3.00 | 0.59 (0.37 – 0.76) | 0.659*** |
| Positive mood (PANAS) (range 2 – 10) | 5.80 (2.09); 6.00 | 0.53 (0.36 – 0.70) | -0.081** | 6.03 (2.01); 6.00 | 0.50 (0.28 – 0.78) | -0.116** |
| Negative mood (PANAS) (range 2 – 10) | 3.91 (1.35); 4.00 | 0.51 (0.33 – 0.68) | 0.265*** | 3.86 (1.94); 3.00 | 0.52 (0.32 – 0.72) | 0.268*** |
| Incontinence anxiety (range: 1 – 5) | 1.86 (1.22); 1.00 | 0.57 (0.37 – 0.74) |  | 2.61 (1.80); 2.00 | 0.78 (0.55 – 0.91) |  |

Note: ICC = Intraclass Correlation Coefficient; ^ $p < .10$ ; \* $p < .05$ ; \*\* $p < .01$ ; \*\*\* $p < .001$

On days with UI, the most common frequency was once during a day (44.8%). The median dry interval was less than four hours, although one-third of UI events were continually (more than four hours) dry. The median (and most common) amount of UI reported was a “medium” amount, while one in five UI days were linked to “large” amount of urine. Nearly all (89.8%) of UI events required no management help, and very few (7.7%) required activity avoidance. The median daily anxiety when UI occurred was relatively low (1.8; scale range: 1.0 – 5.0). None of these variables – daily UI frequency (*p*=0.991), amount (*p*=0.830), dry intervals (*p*=0.671), management (*p*=0.864), avoiding activities (*p*=0.499) and incontinence negativity (*p*=0.973) – occurred more or less frequently by any specific day of the week.

#### Fecal incontinence

As shown in Table 2, YASB reported any FI on a fifth of all days (20.84%).

Converted to days in a week, YASB generally experienced any FI on slightly more than *one out of every seven days in a “typical” week.* Sensitivity analyses suggest that reports of FI were spread relatively evenly across each day of the week (each day 10.4% to 19.9% of all FI reports; *p*=0.055). Reporting FI did not change as a function of study participation progression (e.g., Day 1 vs. Day 13 vs. Day 29).^9^

When FI occurred, it most commonly occurred once per day (82.1%), typically with a small amount of stool leak (52.9%). Half of FI events involved continuous dry intervals (more than four hours) (54.9%). While the majority (88.3%) of FI events were managed independently, one in four events were linked to avoiding activities (26.6%). The median daily anxiety when FI occurred was relatively low (2.6; scale range: 1.0 – 5.0). None of these variables – daily FI frequency (*p*=0.297), amount (*p*=0.485), dry intervals (*p*=0.670), management (*p*=0.740), avoiding activities (*p*=0.703) and FIA (*p*=0.885) – were different by any specific day of the week.

### Objective 1b – Describe Day-to-Day Variation in Prevalence and Characteristics of UI and FI

#### Urinary incontinence

Data suggested more between-person variance (ICC: 0.78 – 0.96) for UI frequency, dry intervals, amount, management, activity avoidance and median past anxiety and more within- person variance (ICC: 0.32 – 0.57) for past UI frequency, positive and negative mood and UI anxiety.

#### Fecal incontinence

We observed more *between-person* variable (ICC: 0.78 – 0.87) for FI management, past FI frequency and current incontinence anxiety, and more within-person variance (ICC: 0.43 – 0.52) for FI frequency, dry intervals, amount, activity avoidance, past FI anxiety and positive and negative mood.

### Objective 2 – Daily Association of UI and FI Prevalence and Characteristics with Incontinence Negativity

The goal of this objective was to explore any daily association between UI or FI prevalence or context and UIA or FIA. In other words, does the way in which a YASB experience a given UI or FI episode impact the extent to which they feel anxiously about that same episode?

Table 2 provides the correlations between daily UI and FI prevalence and characteristics with daily UIA and FIA. *Higher* daily UIA was significantly correlated with increased UI frequency (three or more times vs. once), shorter dry intervals between UI leaks, greater UI leak volume, needing help to manage the leak, avoiding one or more activities because of UI, a higher past median UIA and higher negative mood. *Lower* UIA was significantly correlated with a higher number of UI events in the past.

FIA was linked to fewer daily context variables. *Higher* daily FIA with significantly correlated with shorter dry intervals (less than four hours vs. more than four), a higher past daily median FIA and a higher daily negative mood. *Lower* FIA was significantly correlated with a higher number of UI events in the past.

### Exploratory Objective – Compare the Prevalence, Episode-Specific Characteristics and Incontinence Negativity of UI vs. FI

UI occurred significantly *more* frequently (median: twice vs. once; *p*<.001), with significantly *greater* volume (median: less than four hours vs. continually dry; *p*<.001) and significantly *shorter* dry intervals (median: less than four hours vs. continually dry; *p*<.001) as compared to FI. YASB reported activity avoidance significantly *less often* on days with reports of UI as compared to FI (7.7% vs. 26.7%; *p*<.001). Positive mood was significantly *higher* and negative mood significantly *lower* on days with UI vs. FI (both *p*<.001). YASB felt significantly *less negatively* about experiencing UI than FI (*p*<.001).

## Discussion and Implications

This paper highlighted the first time use of ecological momentary assessment (EMA) to elicit YASB- provided information about daily UI and FI prevalence, episode-specific characteristics and negativity over 30 days. Despite the substantial both short-^6,8,33^ and long-term^7,34^ impact of incontinence in YASB’s lives, broad and retrospective research and clinical measurement of UI and FI often preclude data that actually reflect how YASB live these experiences on a daily basis. Our data provide preliminary indication that the way in which we ask about incontinence matters to accurately understanding how YASB experience UI or FI in their daily lives. While additional research is needed to further refine the best measurement structure and data collection in this population, below we highlight findings that we think are particularly important for incontinence care among YASB.

First, we observed that every metric of UI and FI showed within-person change, that some metrics showed more change than others, and from our supplementary data, that UI and FI were different from one another on the same metrics. Put simply: no two days of UI or FI look the same, and YASB do not experience UI in the same way they experience FI. While both of these statements may seem obvious, they do provide potential points upon which a currently standardized approach to SB incontinence clinical care could be tailored to address individual needs. For example, traditional clinical models typically prioritize treating the most severe of all incontinence symptoms a patient reports,^35,36^ rather than addressing the entirety of a patient’s experience. Our data suggest that daily incontinence experiences are quite nuanced from day-to-day, and that what aspects of UI or FI that YASB find the most bothersome may change over time. A less detailed clinical assessment may not capture these potentially important fluctuations. Asking patients to log pre-clinical visit logs of UI and FI could yield better insight as to which episode-specifics of incontinence occur the most frequently or the most acutely for each patient.^37^ This information could also increase a care team’s ability to design interventions that specifically address a YASB’s unique UI concerns and unique FI concerns, rather than assuming a single intervention will address all concerns.^19^ It will be important for future research to continue exploring the optimal breadth and depth of data collected from patients – as well as the ideal time to review any changes in UI and FI – to support clinical decision making. Our hope is that future work will evaluate how data like these can support flexible YASB-tailored interventions that can be iteratively revised as needed.^38^

Second, the exploratory analyses of daily incontinence negativity revealed several important ideas. Both episode-specific (e.g., activity avoidance) and affective (e.g., feeling in a bad mood) factors increased current day incontinence negativity. While previous work has suggested that different UI and FI factors impact what bothers people about incontinence,^5,15^ our data show that what specifically triggers higher or lower negativity is likely more nuanced than just measuring clinically objective markers of incontinence. In addition, temporal factors – both frequency of past incontinence and past negativity towards incontinence – impacts daily negativity. The idea that our past experiences impact our evaluation of current events is not a new concept in either young adult behavior research,^39–41^ but has yet to be explored in the SB population and warrant further research. For example, a greater number of past UI events predicted *lower* current day UIA, perhaps suggesting that higher past exposure provides a desensitization effect. Higher past UIA and FIA were associated with higher current-day UIA and FIA, potentially reflecting an accumulated “expectation” or routinized negativity. Collectively, these findings – current day episode-specific factors, daily mood and past experiences – are all new points of focus in this population and warrant further investigation in the context of clinical work. While more data are needed, we think these findings promising as just-in-time interventions that could be addressed at the same time incontinence itself is treated. Tailoring real-time solutions within an individual’s own experiences represents a patient-centered approach yet unused in clinical settings.

There are limitations associated with the current data. First, by design, this study excluded individuals under 18 years of age. While our data provide some of the first detailed information about the daily experience of UI and FI among YASB, it is important to recognize the developmental roots of continence self-management in adulthood are likely linked to experiences in adolescence. Future studies may benefit from replicating our assessment in younger SB populations, and/or to follow individuals in this population from adolescent into adulthood to better understand how UI and FI occur. Second, our sample was primarily White, heterosexual, well-educated, and exhibited a high degree of living and mobility independence. There is possibility of self- selection bias within this set of characteristics, meaning that individuals who are more caregiver-dependent, or those with greater degrees of developmental delay, may have been less likely to answer the survey. However, our larger participant pool had reasonably similar gender and neurological characteristics to other samples from SB-focused research.^42–45^ Third, we assessed incontinence negativity on days when UI and FI occurred and did not take into account days without any reports of incontinence. The association of past experiences with current negativity in this study likely supports the investigation the role of anticipatory negativity. Fourth, we captured only 30 days of data from each YASB, which is likely too short of a time period to observe the typical cycle of incontinence and negativity in a person’s life. Future studies will focus on assessing both what an optimal data capture frame might be, and whether this frame should occur in a specific context. Fifth, all participants had a recent history of incontinence. We did not capture whether it occurred per stoma or urethra/anus. We anticipate a more detailed future assessment of a more heterogenous population.

## Conclusion

Our data suggest that daily experiences of UI and FI vary across multiple dimensions – and perhaps more importantly, these dimensions have separate associations with how negatively UI and FI feel about incontinence on a given day. While more data will be needed to further refine our understanding of these relationships, we believe that self-reporting UI and FI experience variability via EMA hold promise to inform a new generation of interventions to improve long-term quality of life in those with SB. Self-monitoring symptoms would support for collaborative patient-clinician diagnosis and evaluation,^38^ ultimately allowing the design of more individually-tailored treatmen^46^ that can be dynamically changed as needed.^47^

## Data Availability

Raw data files and codebooks are stored with the Open Science Framework

https://osf.io/gefqx/

## References

1. Szymanski KM, Roth JD, Hensel DJ, et al. Sexual activity and function of adult men with spina bifida. J Pediatr Urol. Jun 2023;19(3):308 e1-308 e9. doi:10.1016/j.jpurol.2023.03.002

2. Szymanski KM, Misseri R, Whittam B, et al. QUAlity of Life Assessment in Spina bifida for Adults (QUALAS-A): development and international validation of a novel health-related quality of life instrument. Qual Life Res. Oct 2015;24(10):2355–64. doi:10.1007/s11136-015-0988-5

3. Roth JD, Hensel DJ, Wiener JS, et al. Urinary and Fecal Incontinence During Sexual Activity Is Common and Bothersome Among Adults With Spina Bifida. Urology. Apr 2024;186:54–60. doi:10.1016/j.urology.2023.12.029

4. Kelly MS, Wiener JS, Liu T, et al. Neurogenic bowel treatments and continence outcomes in children and adults with myelomeningocele. J Pediatr Rehabil Med. 2020;13(4):685–693. doi:10.3233/prm-190667

5. Szymanski KM, Cain MP, Whittam B, Kaefer M, Rink RC, Misseri R. All incontinence is not created equal: impact of urinary and fecal incontinence on quality of life in adults with spina bifida. The Journal of urology. 2017;197(3):885–891.

6. Page DT, Coetzee BJ. South African adolescents living with spina bifida: contributors and hindrances to well-being. Disability and Rehabilitation. 2021;43(7):920–928.

7. Liu T, Ouyang L, Walker WO, et al. Education and employment as young adults living with spina bifida transition to adulthood in the USA: A study of the National Spina Bifida Patient Registry. Developmental Medicine & Child Neurology. 2023;65(6):821–830.

8. Gabrielsson H, Traav MK, Cronqvist A. Reflections on health of young adults with spina bifida: The contradictory path towards well-being in daily life. Open Journal of Nursing. 2015;5(04):303.

9. Hensel DJ, Young AI, Szymanski KM. The feasibility of using ecological momentary assessment to understand urinary and fecal incontinence experiences in adults with spina bifida: A 30-day study. PLOS ONE. 2023;18(11):e0292735. doi:10.1371/journal.pone.0292735

10. Talari K, Goyal M. Retrospective studies–utility and caveats. Journal of the Royal College of Physicians of Edinburgh. 2020;50(4):398–402.

11. Iida M, Shrout PE, Laurenceau J-P, Bolger N. Using diary methods in psychological research. 2012;

12. Hensel DJ, Fortenberry JD, Harezlak J, Craig D. The Feasibility of Cell Phone Based Electronic Diaries for STI/HIV Research. BMC Medical Research Methodology. 2012;12(75)

13. Roth AM, Hensel DJ, Fortenberry JD, Garfein RS, Gunn JK, Wiehe SE. Feasibility and Acceptability of Cell Phone Diaries to Measure HIV Risk Behavior Among Female Sex Workers. AIDS and Behavior. 2014:1–11.

14. Shiffman S, Stone A, Hufford M. Ecolocial Momentary Assessment. Annual Review of Clinical Psychology. 2008;4:1–32. 10.1146/annurev.clinpsy.3.022806.091415

15. Szymanski KM, Misseri R, Whittam B, Kaefer M, Rink RC, Cain MP. Quantity, not frequency, predicts bother with urinary incontinence and its impact on quality of life in adults with spina bifida. The Journal of urology. 2016;195(4 Part 2):1263–1269.

16. Gabrielsson H, Hultling C, Cronqvist A, Asaba E. Views on everyday life among adults with spina bifida: an exploration through photovoice. International Journal of Qualitative Studies on Health and Well-being. 2020;15(1):1830702.

17. Lim S-W, Yi M. Illness Experiences of Adults with Spina Bifida: Protecting the Whole Self. Asian Nursing Research. 2021/02/01/ 2021;15(1):67-75. 10.1016/j.anr.2020.12.001

18. Szymanski KM, Misseri R, Hensel DJ. Accuracy in reporting incontinence in adults with spina bifida: a pilot study. Journal of Pediatric Urology. 2024;

19. Szymanski KM, Carroll AE, Misseri R, Moore CM, Hawryluk BA, Wiehe SE. A patient-and parent-centered approach to urinary and fecal incontinence in children and adolescents with spina bifida: understanding experiences in the context of other competing care issues. Journal of Pediatric Urology. 2023;19(2):181–189. doi:10.1016/j.jpurol.2022.10.027

20. Joseph NT, Jiang Y, Zilioli S. Momentary emotions and salivary cortisol: A systematic review and meta-analysis of ecological momentary assessment studies. Neuroscience & Biobehavioral Reviews. 2021/06/01/ 2021;125:365–379. 10.1016/j.neubiorev.2021.02.042

21. Hufford M. Special methodological challenges and opportunities in ecological momentary assessment. In: Stone AA, Shiffman S, Atienza A, Nebeling L, eds. The science of real-time data capture: Self-reports in health research Oxford University Press; 2007:54–75.

22. Todd KR, Shaw RB, Kramer JL, Martin Ginis KA. Using ecological momentary assessment to evaluate neuropathic pain experienced by adults with SCI: recommendations and participant perceptions. Disability and Rehabilitation. 2021;43(17):2439–2446.

23. May M, Junghaenel DU, Ono M, Stone AA, Schneider S. Ecological Momentary Assessment Methodology in Chronic Pain Research: A Systematic Review. The Journal of Pain. 2018;

24. Russell MA, Gajos JM. Annual Research Review: Ecological momentary assessment studies in child psychology and psychiatry. Journal of Child Psychology and Psychiatry. 2020;61(3):376–394.

25. Stone AA, Shiffman SS. Ecological validity for patient reported outcomes. In: Steptoe A, ed. Handbook of Behavioral Medicine. Springer; 2010:99–112.

26. Hufford MR, Shiffman S. Methodological issues affecting the value of patient-reported outcomes data. Expert review of pharmacoeconomics & outcomes research. 2002;2(2):119–128.

27. Robbins ML, Kubiak T, Mostofsky D. Ecological momentary assessment in behavioral medicine: Research and practice. The handbook of behavioral medicine. 2014;1:429–446.

28. Struwe S, Thibadeau J, Kelly MS, Widener-Burrows D. Establishing the first community-centered Spina Bifida research agenda. Journal of Pediatric Urology. 2022;18(6):800. e1-800. e7.

29. Van Voorhees EE, Dennis PA, Elbogen EB, et al. Characterizing anger-related affect in individuals with posttraumatic stress disorder using ecological momentary assessment. Psychiatry Research. 2018;261:274–280. doi:10.1016/j.psychres.2017.12.080

30. *Stata*. Version 18.0. StataCorp LP; 2023.

31. Merz EL, Roesch SC. Modeling trait and state variation using multilevel factor analysis with PANAS daily diary data. J Res Pers. Feb 1 2011;45(1):2–9. doi:10.1016/j.jrp.2010.11.003

32. Xiao C, Ye J, Esteves RM, Rong C. Using Spearman’s correlation coefficients for exploratory data analysis on big dataset. Concurrency and Computation: Practice and Experience. 2016;28(14):3866–3878.

33. Ridosh MM, Sawin KJ, Roux G, Brei TJ. Quality of life in adolescents and young adults with and without spina bifida: an exploratory analysis. Journal of Pediatric Nursing. 2019;49:10–17.

34. Liptak GS, Kennedy JA, Dosa NP. Youth with Spina Bifida and Transitions: Health and Social Participation in a Nationally Represented Sample. The Journal of Pediatrics. 2010/10/01/ 2010;157(4):584–588.e1. 10.1016/j.jpeds.2010.04.004

35. Pasch L, He SY, Huddleston H, et al. Clinician vs Self-ratings of Hirsutism in Patients With Polycystic Ovarian Syndrome: Associations With Quality of Life and Depression. JAMA dermatology. Mar 4 2016;doi:10.1001/jamadermatol.2016.0358

36. Hinami K, Alkhalil A, Chouksey S, Chua J, Trick WE. Clinical significance of physical symptom severity in standardized assessments of patient reported outcomes. Qual Life Res. Mar 15 2016;doi:10.1007/s11136-016-1261-2

37. Piot M, Mestdagh M, Riese H, et al. Practitioner and researcher perspectives on the utility of ecological momentary assessment in mental health care: A survey study. Internet Interventions. 2022/12/01/ 2022;30:100575. 10.1016/j.invent.2022.100575

38. van Os J, Verhagen S, Marsman A, et al. The experience sampling method as an mHealth tool to support self-monitoring, self-insight, and personalized health care in clinical practice. Depression and anxiety. 2017;34(6):481–493.

39. Hensel DJ, Fortenberry JD, Orr DP. Variations in Coital and Noncoital Sexual Repertoire among Adolescent Women. doi: DOI: 10.1016/j.jadohealth.2007.07.009. Journal of Adolescent Health. 2008;42(2):170–176.

40. Hensel DJ, Fortenberry JD, Orr DP. Factors Associated with Event Level Anal Sex and Condom Use During Anal Sex Among Adolescent Women. doi: DOI: 10.1016/j.jadohealth.2009.06.025. Journal of Adolescent Health. 2010;46(3):232–237.

41. Hensel DJ, Stupiansky NW, Orr DP, Fortenberry JD. Event-Level Marijuana Use, Alcohol Use, and Condom Use Among Adolescent Women. Sexually Transmitted Diseases. 2011;38(3):239–243 doi:10.1097/OLQ.0b013e3181f422ce

42. Szymanski KM, Misseri R, Whittam B, et al. QUAlity of Life Assessment in Spina bifida for Adults (QUALAS-A): development and international validation of a novel health-related quality of life instrument. Quality of Life Research. 2015;24(10):2355–2364.

43. Lemelle J, Guillemin F, Aubert D, et al. Quality of life and continence in patients with spina bifida. Quality of Life Research. 2006;15(9):1481–1492.

44. Schechter MS, Liu T, Soe M, Swanson M, Ward E, Thibadeau J. Sociodemographic attributes and spina bifida outcomes. Pediatrics. 2015;135(4):e957–e964.

45. Sawin KJ, Liu T, Ward E, et al. The National Spina Bifida Patient Registry: profile of a large cohort of participants from the first 10 clinics. The Journal of pediatrics. 2015;166(2):444–450. e1.

46. McKeon A, McCue M, Skidmore E, Schein M, Kulzer J. Ecological momentary assessment for rehabilitation of chronic illness and disability. Disability and rehabilitation. 2018;40(8):974–987.

47. Schwartz S, Schultz S, Reider A, Saunders EF. Daily mood monitoring of symptoms using smartphones in bipolar disorder: a pilot study assessing the feasibility of ecological momentary assessment. Journal of affective disorders. 2016;191:88–93.

