## Supplemental Material for "Daily Experiences of Urinary and Fecal Incontinence in Young Adults with Spina Bifida: Preliminary Results from an Ecological Momentary Assessment Study"

**Supplemental Results**

*Person Level Variation in UI*. This section describes between-person summaries of UI information during the study. All data refer to information *per person.*

As shown in Supplemental Table 1, the majority (95.6%; N=22) of YASB reported any UI during the 30-day period. The number of days with UI ranged from zero days to all 30 days, with median of 9.0 days (Table 2). In other words, while the “typical” YASB reported UI on about half of the days out of every month, these reports varied across all YASB (Supplemental Figure 1).

Across all reports of UI during the study, about half of YASB (59.1%) reported that UI occurred a median of once per day and with a median medium volume (50.0%). An additional half of YASB noted they were continually dry following UI events. The median number of UI events (eight out of nine median UI events). Although most UI events were managed independently (median: eight events out of a median nine total UI events reported), the total number ranged from zero to 30. Activities were missed on a median of zero (out of nine median) UI days (range: 0 – 14).

The YASB-level median UIN score was 6.7 (out of 20) per UI event, although more than half (54.5%) of YASB reported a median of 7.0 or less. A third of YASB reported a median no bother (minimum value: four) and only five percent reported a median highest bother possible (maximum value: 20). In other words, while the “average” YASB reported a middle level of UIN, the value varied across all YASB (Supplemental Figure 2).

*Person Level Variation in FI.* This section describes between-person summaries of FI information during the study. All data refer to information *per person.*

As shown in Supplementary Table 1, three-quarters (73.9%; N=17) of YASB reported any FI during the 30-day period. The number of days with FI ranged from zero days to 29 days, with mean of almost six days (median: six days). In other words, while the “average” YASB reported FI on about six of the days out of every month, the frequency varied across all YASB (Supplemental Figure 1).

Across all reports of FI during the study, the majority (78.3%) reported that FI occurred a median of once per day and about half (47.1%). An additional half of YASB (56.1%) noted they were continually dry following UI events. Although about all FI events were managed independently (median: three events out of a median three total FI events over the 30 days), the total number ranged from zero to 30. Activities were missed on a median total of three (out of nine median) FI days (range: 0 – 14).

The YASB-level median FIN score was 9.0 (out of 20) per FI event, although more than half (54.5%) of YASB reported a median of 7.0 or less. One quarter of YASB reported a median no bother (minimum value: four) and twenty percent reported a median highest bother possible (maximum value: 20). Nearly half of the sample suggested a median FIN at the midpoint (10.0) or higher. In other words, while the "average" YASB reported a middle level of FIN, the value varied across all YASB (Supplemental Figure 2).

Supplemental Table 1. Person-Level Summary of Urinary (UI) and Fecal Incontinence (FI) Prevalence and Context Over 30 Days among Young Adults with Spina Bifida (N=23).

| Incontinence Characteristic | Percent or Mean (SD); Median |
|---|---|
| Any UI during study (yes) | 95.6 |
| Any FI during study (yes) | 73.9 |
| Any incontinence during study (yes) | 95.6 |
| UI only | 21.7 |
| FI only | 4.3 |
| Both UI and FI | 73.9 |
| Total number of days with incontinence during study | |
| Urinary (range: 0 – 30) | 15.3 (12.1); 9.0 |
| Fecal (range: 0 – 29) | 5.8 (8.1); 3.0 |
| Median Incontinence Negativity during study | |
| Urinary (UIN; range: 4 – 20) | 7.7 (4.6); 6.0 |
| Fecal (FIN; range: 4 – 20) | 11.2 (6.33); 9.0 |
| Median UI frequency during study | |
| Once | 59.1 |
| Twice | 18.2 |
| Three or more times | 22.7 |
| Median FI frequency during study | |
| Once | 82.4 |
| Twice | 17.6 |
| Three or more times | 0.0 |
| Median UI volume during study | |
| Small amount | 36.4 |
| Medium amount | 50.0 |
| Large amount | 13.6 |
| Median FI volume during study | |
| Small amount | 41.2 |
| Medium amount | 41.2 |
| Large amount | 17.7 |
| Median UI dry intervals during study | |
| More than four hours | 50.0 |
| Less than four hours | 45.5 |
| Never | 4.5 |
| Median FI dry intervals during study | |
| More than four hours | 0.0 |
| Less than four hours | 43.8 |
| Never | 56.3 |
| Total number of independently managed incontinence events during study | 13.2 (11.5); 8.0 |
| Urinary (range: 0 - 30) | 5.4 (8.4); 5.0 |
| Fecal (range: 0 - 29) | |
| Total number of days of missed activities due to incontinence during study | 1.4 (3.4); 0.0 |
| Urinary (range: 0 – 14) | 1.6 (2.7); 3.0 |
| Fecal (range: 0 – 11) | |

Supplemental Figure 1. Person Level Reports of Total Daily Urinary and Incontinence Reports and Median Incontinence Negativity over 30 Days among Young Adults with Spina Bifida.

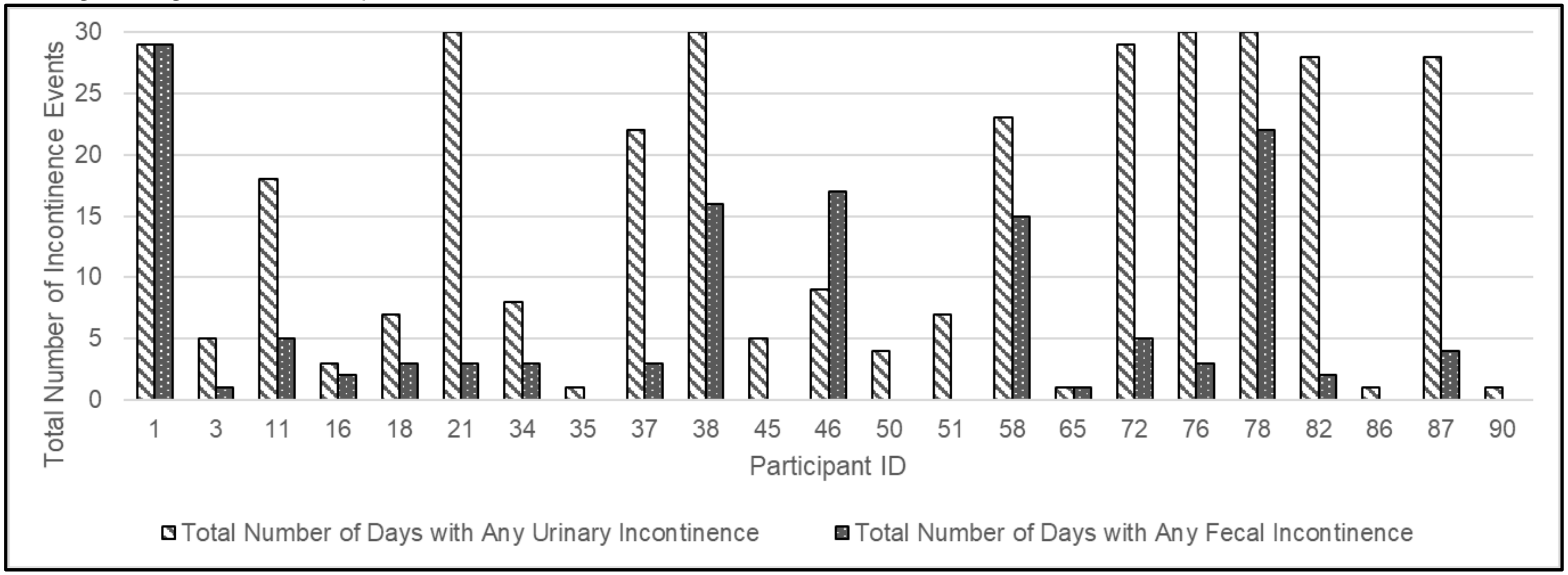


Supplemental Figure 2. Person Level Reports of Median Urinary Incontinence Negativity (UIN) and Fecal Incontinence Negativity (FIN) over 30 Days among Young Adults with Spina Bifida.

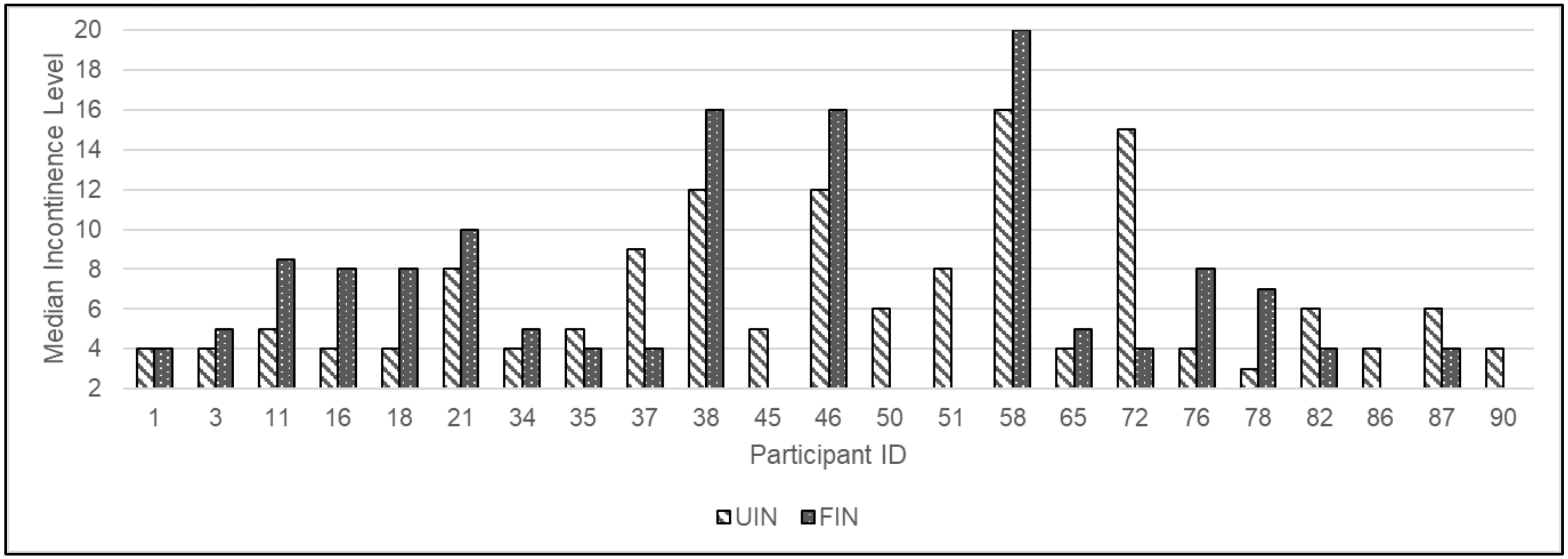
